# Accelerate aging meditates association between bowel dysfunction and depression severity

**DOI:** 10.64898/2026.02.04.26345571

**Authors:** Genghui Zhang, Xuning Wang, Xinyu Wang, Cheng Zhang

## Abstract

**Background:** Our study aimed to investigate the relationship between phenotypic age acceleration (PAA), bowel dysfunction (constipation, diarrhea), and depression severity, and examine whether phenotypic age acceleration can play a mediating role in bowel dysfunction and depression severity.

**Methods:** The data analysis of our study was conducted from the National Health and Nutrition Examination Survey (2005–2010). Participants with bowel dysfunction were identified on the questionnaire of bowel health. Depression was determined based on the Patient Health Questionnaire-9 (PHQ-9). The calculation of PAA is based on 9 test indicators and actual age; a higher PAA means accelerated aging. In this study, a weighted linear regression model was used to analyze the associations among defecation disorders, PAA, and depression. Restricted Cubic Spline (RCS) curves were applied to explore the potential non-linear relationships between the aforementioned variables. Additionally, a mediation effect model was employed to verify whether PAA could function as a mediating variable in the relationship between defecation disorders and depression.

**Result:** A total of 11,808 participants were included in this study. Linear regression analysis showed that both diarrhea (β=3.73, 95% Confidence Interval (CI): 1.69-8.22, P=1.60×10-3) and depression severity (β=1.08, 95%CI: 1.06-1.09, P=4.61×10-16) were positively correlated with PAA. In addition, both constipation (β=2.76, 95%CI: 1.89-4.04, P=2.28×10^−6^) and diarrhea (β=4.29, 95%CI: 2.65-6.95, P=2.11×10-7) were positively correlated with depression severity. Further mediation effect analysis revealed that PAA may play a mediating role in the association between diarrhea and depression severity (the proportion of mediation effect in the total population was 7.2285%). When exploring whether PAA exerts a mediating role in the association between constipation and depression severity, it was found that PAA played a mediating role in female participants and participants aged <60 years, except for male participants and those aged ≥60 years (the proportion of mediation effect was 9.8417% in females and 8.4512% in the population aged <60 years, with all relevant P-values <0.005)

## 1. Introduction

Constipation and diarrhea are categorized as functional gastrointestinal disorders (FGIDs), which are primarily characterized by changes in stool form and defecation frequency[1]. These conditions affect approximately 2.9% to 4.1% of the global population[2]. During the occurrence and progression of the diseases[3], they are often accompanied by psychological factors such as mental stress, anxiety, and depression. Besides reducing patients’ quality of life, these disorders also impose a negative economic burden on the global healthcare system.

Aging is a complex systemic process driven by the gradual accumulation of various molecular and cellular damages. When the rate of this process accelerates, it is termed “accelerated aging,” which is often accompanied by an increased risk of chronic diseases and degenerative changes in the body. Previous studies have confirmed that intestinal flora imbalance caused by constipation and diarrhea is a key factor driving the aging process[4], [5], and it may promote the occurrence and progression of aging through pathways such as inducing chronic inflammation and regulating metabolites[6].

Phenotypic age (PA) is a type of biological age calculated based on blood biomarkers, whose core purpose is to measure the degree of an individual’s physiological aging. Currently, its role in clinical risk assessment, public health research, and anti-aging intervention trials has been fully verified[7], [8], [9].

Among the various factors that promote aging, in addition to intestinal flora, the role of depression has also been widely confirmed. Depression can accelerate the aging process by affecting the Frailty Index (FI), Telomere Length (TL), and Appendicular Lean Mass (ALM), and this aging-promoting effect has also been verified in studies related to phenotypic age. Furthermore, previous studies have shown that there is a bidirectional interaction between depression and functional gastrointestinal disorders: on one hand, depressive states may increase the risk of developing functional gastrointestinal disorders; on the other hand, intestinal flora imbalance caused by constipation and diarrhea may also induce depression by affecting the microbiota-gut-brain axis and disrupting the function of the central nervous system.

Considering the interactions among aging, FGIDs, and depression, we propose the following hypothesis: aging may modulate the occurrence and progression of FGIDs, which in turn exacerbates the severity of depression. To verify this conjecture, we plan to collect and analyze relevant data from the National Health and Nutrition Examination Survey (NHANES) to clarify the correlation between FGIDs and depression, and evaluate the potential mediating role of PAA in regulating the severity of depression.

## 2. Methods

### 2.1 Study population

The NHANES is a major program run by the U.S. Centers for Disease Control and Prevention that aims to collect comprehensive data on the health and nutritional status of the U.S., which encompasses multi-dimensional data, including demographics, nutrition, physical examinations, laboratory tests, and questionnaires. It supports research on a wide range of diseases and health conditions, such as malignant tumors and cardiovascular diseases. Additionally, its data can be used to analyze the pathogenesis and epidemiological trends of diseases, thereby advancing research and development across various fields of medicine.

Participants from the NHANES cycles spanning 2005 to 2010 were included in this study. Firstly, we excluded participants under the age of 20 (N=13902). Secondly, participants with missing data on depression and other covariates were excluded (N=5324). Finally, a total of 11808 participants were included in the study, consisting of 882 patients with constipation and 891 patients with diarrhea. Figure 1 illustrates the process of participant screening.

**Figure 1.**
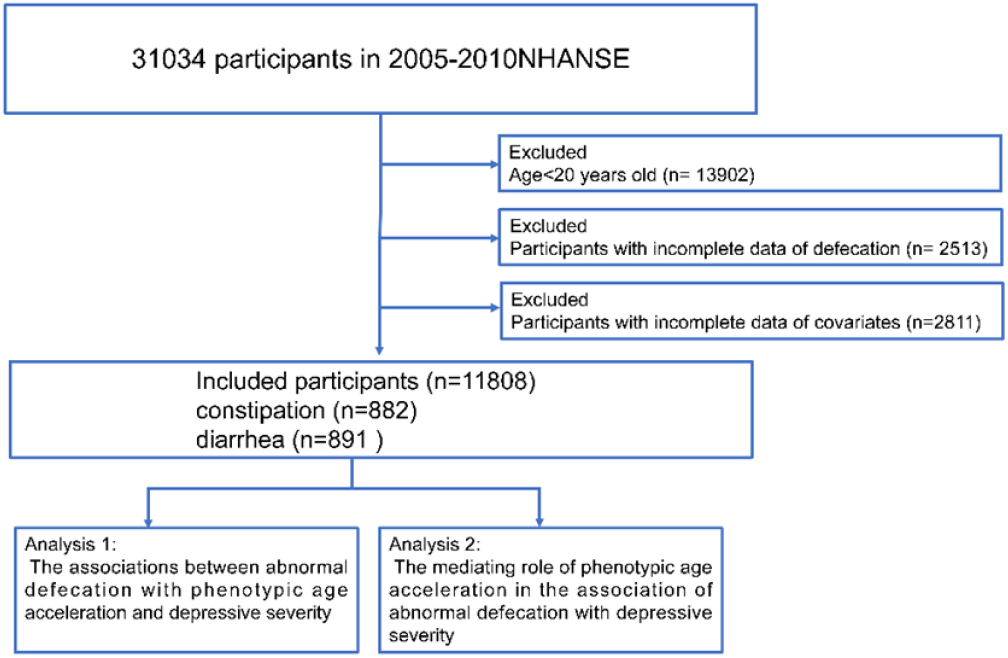
Flow chart of participant selection. NHANES

### 2.2 FGIDs Status

FGIDs status was assessed via a questionnaire, which administered in person in the MEC interview room to survey participants only: “Please look at this card and tell me the number that corresponds to your usual or most common stool type.”. participants’ responses were restricted to the following categories: Type 1, defined as separate hard lumps like nuts (corresponding to constipated stool); Type 2, described as sausage-like but lumpy (corresponding to mild constipated stool); Type 3, which is sausage-like but with cracks on the surface (corresponding to stool close to normal); Type 4, characterized as sausage-like or snake-like, smooth and soft (corresponding to normal stool); Type 5, referring to soft blobs with clear-cut edges (corresponding to mild diarrheal stool); Type 6, meaning fluffy pieces with ragged edges and presenting as mushy stool (corresponding to moderate diarrheal stool); and Type 7, defined as watery with no solid pieces (corresponding to severe diarrheal stool). Additionally, two non-substantive response options were provided for participants: “Refused” (indicating refusal to answer) and “Don’t know” (indicating inability to determine the category).

In this study, according to the Bristol Stool Scale[10], participants who reported their stool characteristics as Type 1 (separate hard lumps, like nuts) and Type 2 (sausage-like but lumpy) were defined as the constipation group; those who reported Type 6 (fluffy pieces with ragged edges, mushy stool) and Type 7 (watery, no solid pieces) were defined as the diarrhea group. After excluding participants who selected “Refused” and “Don’t know”, the remaining participants who reported other stool types (i.e., Type 3, Type 4, and Type 5) were defined as the normal group.

### 2.3 Depression severity

Depressive symptoms in participants were assessed using the PHQ-9 questionnaire (Patient Health Questionnaire-9). Participants were asked to recall and report their conditions over the past two weeks in accordance with the questionnaire instructions. Widely used internationally for evaluating the severity of depression, this questionnaire quantifies depressive severity by scoring 9 core depression-related symptoms (each symptom is scored on a 0-3 scale, with a total possible score ranging from 0 to 27). Participants with a total score of 10 or higher were defined as having “depression”; Those with a total score of less than 10 were determined to have “no clinically relevant depressive symptoms”[11].

### 2.4 phenotypic age acceleration

PA is a biological age indicator calculated based on blood biomarkers. It assesses an individual’s degree of biological aging by measuring the functional status of multiple physiological systems in the human body. Through a multiple regression model, it correlates nine clinically commonly used biomarkers—including creatinine level, albumin level, the logarithm of C-reactive protein (CRP) level, glucose level, alkaline phosphatase level, lymphocyte percentage, red blood cell distribution width (RDW), mean corpuscular volume (MCV), and white blood cell (WBC) count—with mortality risk, and finally outputs a value expressed as “age”[12] .

PA is a more accurate indicator for predicting health risks than chronological age, and can serve as an effective quantitative index for assessing the process of biological aging. Its calculation formula is as follows: PA = 141.50 + (ln [− 0.00553*ln (1-mortality risk)])/0.09165, where mortality risk = 1 - exp. (− 1.51714*exp.(xb)/0.0076927); and xb = − 19.907–0.0336*albumin +0.0095*creatine +0.0195*glucose +0.0954*ln (CRP) – 0.0120*lymphocyte percentage + 0.0268*mean cell volume + 0.3356*red blood cell distribution width + 0.00188*alkaline phosphatase +0.0554*white blood cell count +0.0804*chronological age.

PAA is an index calculated from the linear regression residuals of PA and chronological age. This indicator can directly reflect an individual’s actual aging status: if the PAA value is > 0, it indicates that the individual is in a state of accelerated aging compared with peers; if the PAA value is < 0, it means the individual’s aging rate is slower than that of peers, i.e., in a state of decelerated aging.

### 2.5 Covariate

The covariates of our study included age, gender, race, educational level, marital status, the family poverty income ratio (PIR), BMI, waist circumference, smoking, and drinking. Except for smoking and drinking, data on all other covariates are obtained through demographic questionnaires. Based on the criteria of whether the total number of cigarettes smoking exceeded 100 and whether they are currently smoking, divided the participants into three categories: Never smoking (the total number of cigarettes smoked is less than 100 in their lifetime), former smoking (the total number of cigarettes smoked is less than 100 but have since quit), and current smoking (the total number of cigarettes smoked is less than 100 in their lifetime and continue to smoke presently).

Alcohol consumption status is defined based on questionnaire survey results: using the question “Had at least 12 alcohol drinks/1 yr?” as the criterion, participants who answered “Yes” are classified as “alcohol consumers”.

### 2.6 Statistical analysis

All analyses accounted for the complex, multistage, clustered sampling design of the NHANES to generate estimates that are nationally representative of the non-institutionalized US civilian population. We combined three survey cycles (2005-2006, 2007-2008, and 2009-2010) and created new adjusted sample weights for the combined dataset according to the analytical guidelines provided by the National Center for Health Statistics (NCHS). Specifically, the original mobile examination center (MEC) weights from each two-year cycle were divided by 3 to create the new six-year weight.

The primary sampling unit (SDMVPSU) and stratum (SDMVSTRA) variables were incorporated into all models to accurately compute standard errors and confidence intervals. Descriptive statistics are presented as weighted means (± standard errors) for continuous variables and weighted percentages (± standard errors) for categorical variables.

To verify the reliability of the research conclusions, a multivariate model construction strategy was adopted throughout the data analysis process of this study. Among them, the grouping basis was not adjusted during the subgroup analysis, while the scope of variable adjustment remained consistent across all other models. The specific adjustment was implemented through three progressively layered models. Model 1: No adjustment variables were included. Model 2: demographic variables were incorporated for adjustment, specifically including age, gender, ethnicity, educational level, PIR, and marital status; Model 3: Based on Model 2, lifestyle and anthropometric variables were further included for adjustment, with the newly added variables being smoking status, alcohol consumption, BMI, and waist circumference.

Univariate and multivariate regression models were used to explore the correlations between PAA and constipation (or diarrhea), between PAA and depression severity, and between constipation (or diarrhea) and depression severity. RCS curves were applied to visualize the non-linear relationships between PAA and constipation (or diarrhea), as well as between PAA and depression severity. Mediation effect analysis was conducted to clarify whether PAA plays a mediating role in the association between constipation (or diarrhea) and depression. Additionally, subgroup analyses were performed by gender and age groups (< 60 years old, ≥ 60 years old).

All data analyses were performed using R software (Version 4.51), and a P-value < 0.05 was considered statistically significant.

## 3. Result

### 3.1 Demographic characteristics

The demographic characteristics of participants classified by FGIDs status and depressive status are as follows (Table 1): Compared with normal participants, those in the diarrhea group were older (mean age: 52.9 years vs. 48.9 years), had a higher proportion of females (57.7% vs. 48.1%), a lower proportion of non-Hispanic whites (45.2% vs. 52.3%), a higher proportion of widowed/divorced individuals (26.9% vs. 21.1%), and lower educational attainment and income levels (all P < 0.05); those in the constipation group were younger (mean age: 46.6 years vs. 48.9 years), had a higher proportion of females (68.9% vs. 48.1%), a lower proportion of non-Hispanic whites (44.9% vs. 52.3%), a higher proportion of widowed/divorced individuals (24.1% vs. 21.1%), and lower educational attainment and income levels (all P < 0.05).

**Table 1.**
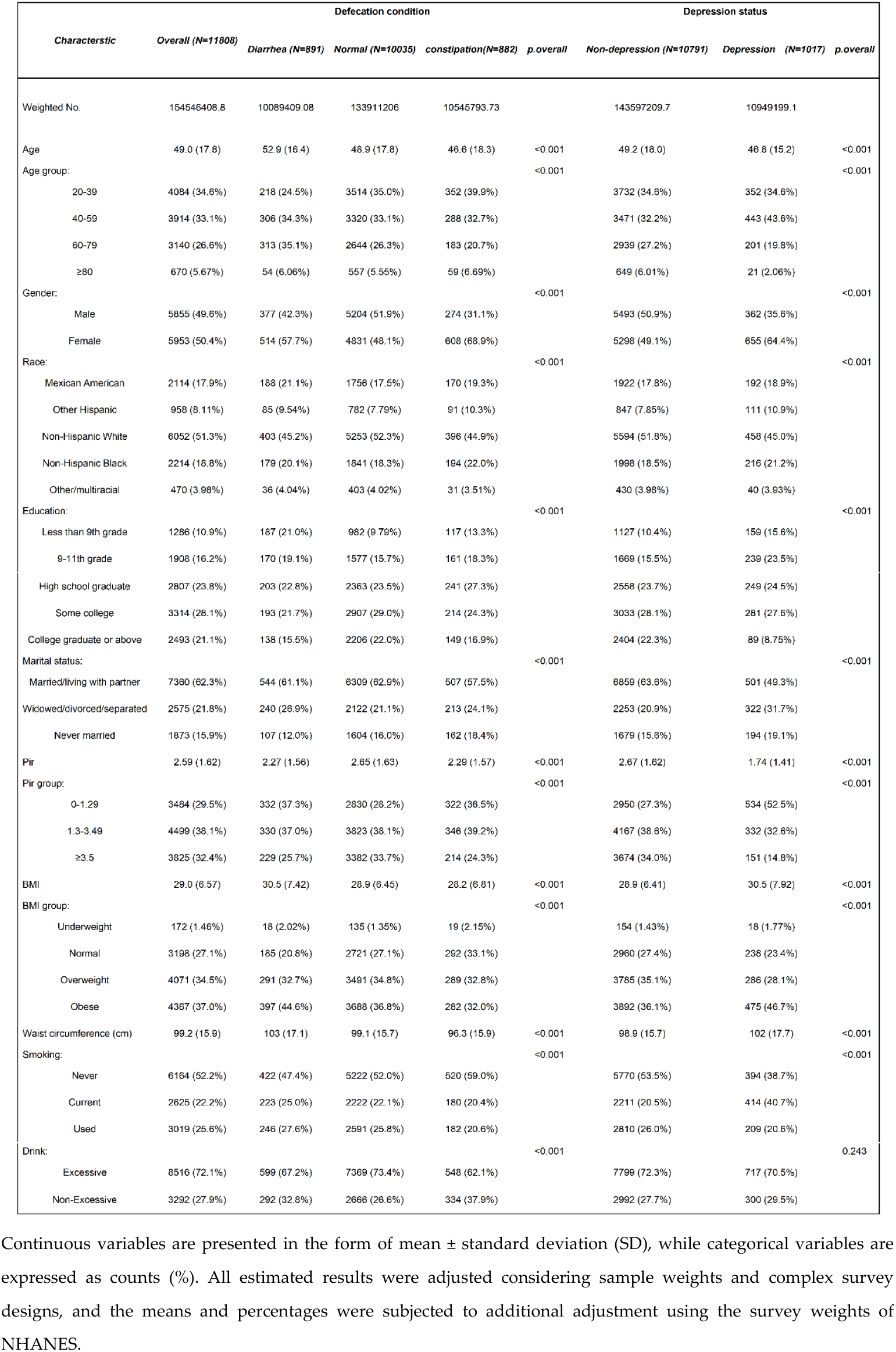
Basic characteristics of the participants included on the basis of FGIDs status and depression status.

Similarly, compared with non-depressive individuals, participants in the depressive group were younger (mean age: 41.64 years vs. 43.54 years), had a higher proportion of females (64.4% vs. 49.1%), a lower proportion of non-Hispanic whites (45.0% vs. 51.8%), a higher proportion of unmarried individuals (31.7% vs. 20.6%), and a lower socioeconomic status (1.65 vs. 1.61), with all the above differences being statistically significant (P < 0.05).

Figure 2 shows histograms demonstrating the distributions of PAA (panels A), depression severity scores and PAA in participants with constipation or diarrhea compared with normal participants (panels B and C), and PAA in patients with different severities of depression (panels D).

**Figure 2.**
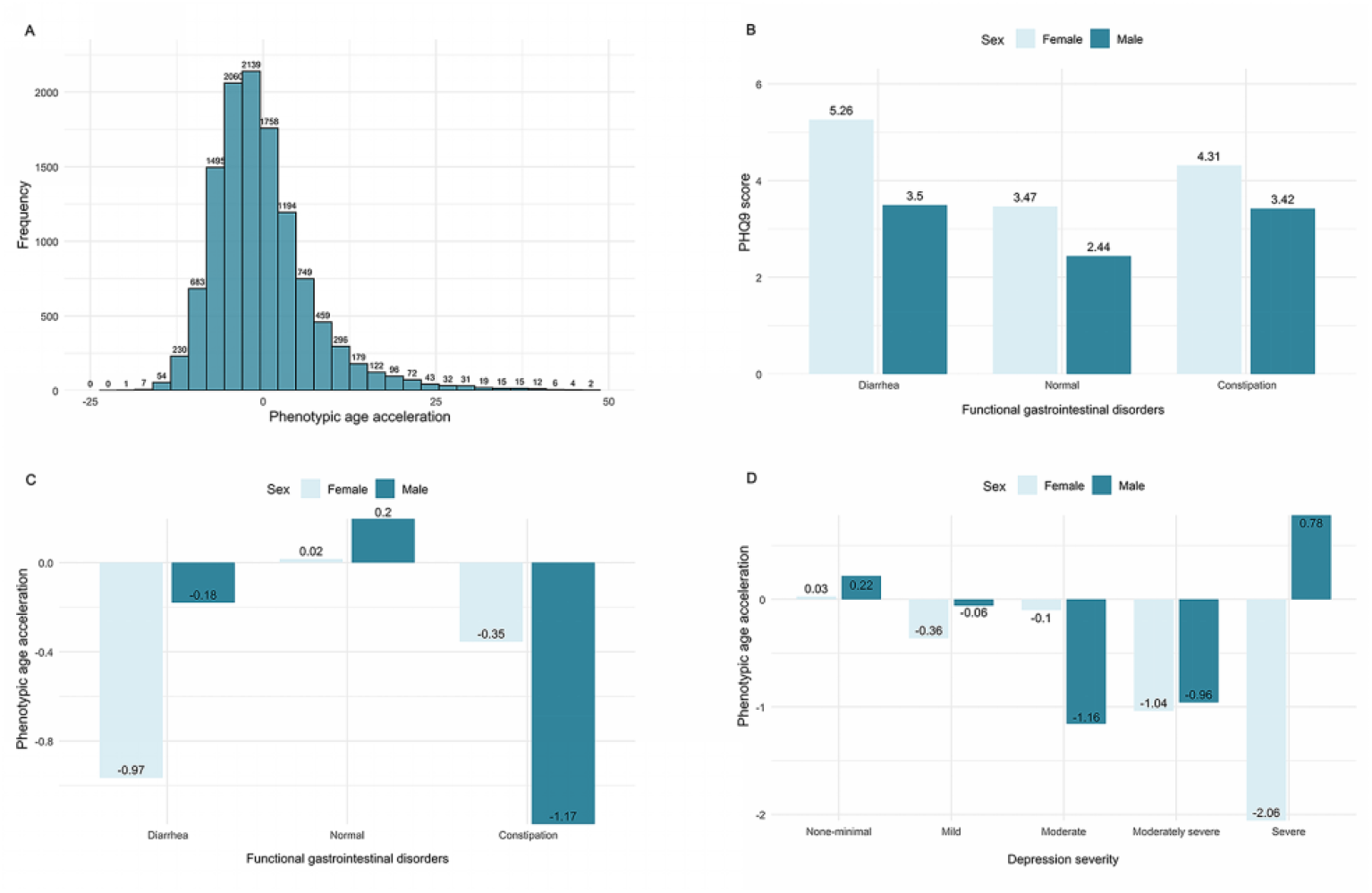
Distributions of phenotypic age acceleration (PAA) and Patient Health Questionnaire-9 (PHQ-9) depression severity scores. A shows the distributions of PAA, B and C display the histograms of PHQ-9 scores and PAA in the overall sample by FIGIDs status. D. Histograms of PAA in participants with different severities of depression

### 3.2. Associations of FGIDs status with PAA and depression severity

In this study, linear regression analysis was employed to explore the correlations between PAA and constipation (or diarrhea), between PAA and depression, and between constipation (or diarrhea) and depression (Table 2). After adjusting for confounding factors, analyses of the total population and subgroups revealed that the correlation between PAA and diarrhea was significantly stronger than that between PAA and constipation. Specifically, in all models of the total population and all models of subgroup analyses, more P-values from the correlation analysis of PAA and diarrhea reached the level of statistical significance. Additionally, the association trends of the two pairs (PAA with diarrhea, PAA with constipation) were similar, both showing the characteristic that “the β-values of female participants and participants aged < 60 years were larger and statistically significant”.

**Table 2.** Linear regression of the associations between diarrhea, constipation, phenotypic age acceleration, and depressive severity.

| Characteristic | Model1 |  |  | Model2 |  |  | Model3 |  |  |
| --- | --- | --- | --- | --- | --- | --- | --- | --- | --- |
| | $\beta$ | 95%CI | P-value | $\beta$ | 95%CI | P-value | $\beta$ | 95%CI | P-value |
| diarrhea and PAA |  |  |  |  |  |  |  |  |  |
| All participants | 3.73 | 1.69-8.22 | 1.60e- 3 | 2.93 | 1.42-6.09 | 5.10e- 3 | 1.57 | 0.807-3.06 | 1.74e- 1 |
| Male | 2.61 | 0.838-8.15 | 0.0958 | 1.93 | 0.661-5.63 | 0.221 | 1.39 | 0.471-4.07 | 0.54 |
| Female | 5.82 | 2.19-15.5 | 7.29e- 4 | 3.68 | 1.39-9.72 | 0.0103 | 1.65 | 0.723-3.78 | 2.22e- 1 |
| <60 | 5.38 | 1.97-14.7 | 1.54e- 3 | 4.32 | 1.71-10.9 | 0.003 | 2.32 | 1.00-5.35 | 4.90e- 2 |
| $\geq 60$ | 1.45 | 0.524-4.00 | 0.468 | 1.13 | 0.364-3.51 | 8.27e- 1 | 0.632 | 0.229-1.75 | 0.361 |
| diarrhea and depression severity |  |  |  |  |  |  |  |  |  |
| All participants | 4.29 | 2.65-6.95 | 2.11e- 7 | 3.56 | 2.24-5.66 | 3.52e- 6 | 3.09 | 1.95-4.91 | 3.38e- 5 |
| Male | 2.44 | 1.32-4.52 | 5.55e- 3 | 2.55 | 1.41-4.60 | 2.93e- 3 | 2.39 | 1.30-4.37 | 0.00665 |
| Female | 5.65 | 2.78-11.5 | 1.17e- 5 | 4.41 | 2.28-8.53 | 6.50e- 5 | 3.61 | 1.89-6.91 | 0.000396 |
| <60 | 4.96 | 2.81-8.77 | 9.09e- 7 | 3.85 | 2.27-6.53 | 1.15e- 5 | 3.38 | 1.96-5.82 | 1.03e- 4 |
| $\geq 60$ | 3.54 | 1.71-7.32 | 1.07e- 3 | 2.68 | 1.35-5.33 | 0.00639 | 2.43 | 1.22-4.85 | 0.0137 |
| constipation and PAA |  |  |  |  |  |  |  |  |  |
| All participants | 1.66 | 0.793-3.48 | 1.74e- 1 | 1.44 | 0.662-3.13 | 3.46e- 1 | 1.93 | 0.966-3.86 | 6.15e- 2 |
| Male | 2.14 | 0.821-5.59 | 0.116 | 1.3 | 0.524-3.24 | 0.557 | 1.55 | 0.666-3.61 | 2.96e- 1 |
| Female | 2.18 | 0.888-5.35 | 8.76e- 2 | 1.43 | 0.573-3.59 | 0.43 | 2.1 | 0.935-4.72 | 7.08e- 2 |
| <60 | 2.62 | 1.17-5.88 | 2.03e- 2 | 1.93 | 0.839-4.46 | 0.118 | 2.47 | 1.22-5.01 | 0.0139 |
| $\geq 60$ | 0.28 | 0.0728-1.08 | 0.0632 | 0.475 | 0.132-1.71 | 2.45e- 1 | 0.761 | 0.220-2.63 | 0.655 |
| constipation and depression severity |  |  |  |  |  |  |  |  |  |
| All participants | 2.76 | 1.89-4.04 | 2.28e- 6 | 1.86 | 1.31-2.64 | 1.09e- 3 | 2.04 | 1.44-2.89 | 2.90e- 4 |
| Male | 2.93 | 1.48-5.78 | 2.64e- 3 | 2.24 | 1.16-4.30 | 1.72e- 2 | 2.33 | 1.22-4.46 | 0.0124 |
| Female | 2.04 | 1.29-3.21 | 2.89e- 3 | 1.68 | 1.09-2.59 | 2.12e- 2 | 1.92 | 1.24-3.00 | 0.00538 |
| <60 | 2.67 | 1.68-4.27 | 1.09e- 4 | 1.79 | 1.16-2.76 | 9.95E-03 | 1.96 | 1.28-3.01 | 3.14e- 3 |
| $\geq 60$ | 2.92 | 1.49-5.72 | 2.45e- 3 | 2.14 | 1.11-4.13 | 0.0249 | 2.31 | 1.19-4.50 | 0.0159 |
| PAA and depression severity |  |  |  |  |  |  |  |  |  |
| All participants | 1.08 | 1.06-1.09 | 4.61E-16 | 1.06 | 1.05-1.08 | 5.03E-11 | 1.04 | 1.03-1.06 | 3.47e- 7 |
| Male | 1.07 | 1.05-1.09 | 3.40e- 8 | 1.06 | 1.03-1.08 | 1.57e- 5 | 1.05 | 1.02-1.07 | 0.000215 |
| Female | 1.1 | 1.07-1.12 | 2.06E-11 | 1.06 | 1.04-1.09 | 1.63e- 7 | 1.04 | 1.02-1.06 | 0.00151 |
| <60 | 1.09 | 1.07-1.11 | 7.05E-12 | 1.06 | 1.04-1.08 | 1.13e- 7 | 1.04 | 1.02-1.06 | 1.03e- 4 |
| $\geq 60$ | 1.05 | 1.03-1.07 | 1.85e- 6 | 1.06 | 1.04-1.08 | 4.16E-06 | 1.05 | 1.03-1.07 | 0.000126 |

Model 1: Crude
Model 2: Adjusted for age, gender, race, pir, education and marital status
Model 3: Model2 + BMI, waist circumference, smoking status and alcohol intake,
Gender was not adjusted when conducting subgroup analyses for males of females
Defecation condition was adjusted when conducting subgroup analyses for PAA and depression severity.

In the correlation analysis with depression, the associations of PAA, constipation, and diarrhea with depression were all statistically significant both before and after variable adjustment. Among them, the correlation between diarrhea and depression was the strongest (β-value comparison: diarrhea 3.09 vs PAA 1.93 vs constipation 1.04).

To further investigate whether there is a non-linear relationship between PAA and the above three factors, we conducted a restricted cubic spline (RCS) analysis with three knots (Figure 3). The results showed that: in the crude model without adjusting for variables, PAA had a significant overall association with all three factors (P overall < 0.05); however, a significant non-linear association was only detected in the analysis of PAA and depression (P non-linear < 0.05), and this association trend was consistent between male and female groups. After variable adjustment, only a significant overall association remained between PAA and the three factors (P overall < 0.05), while no non-linear associations were detected among the three factors.

**Figure 3.**
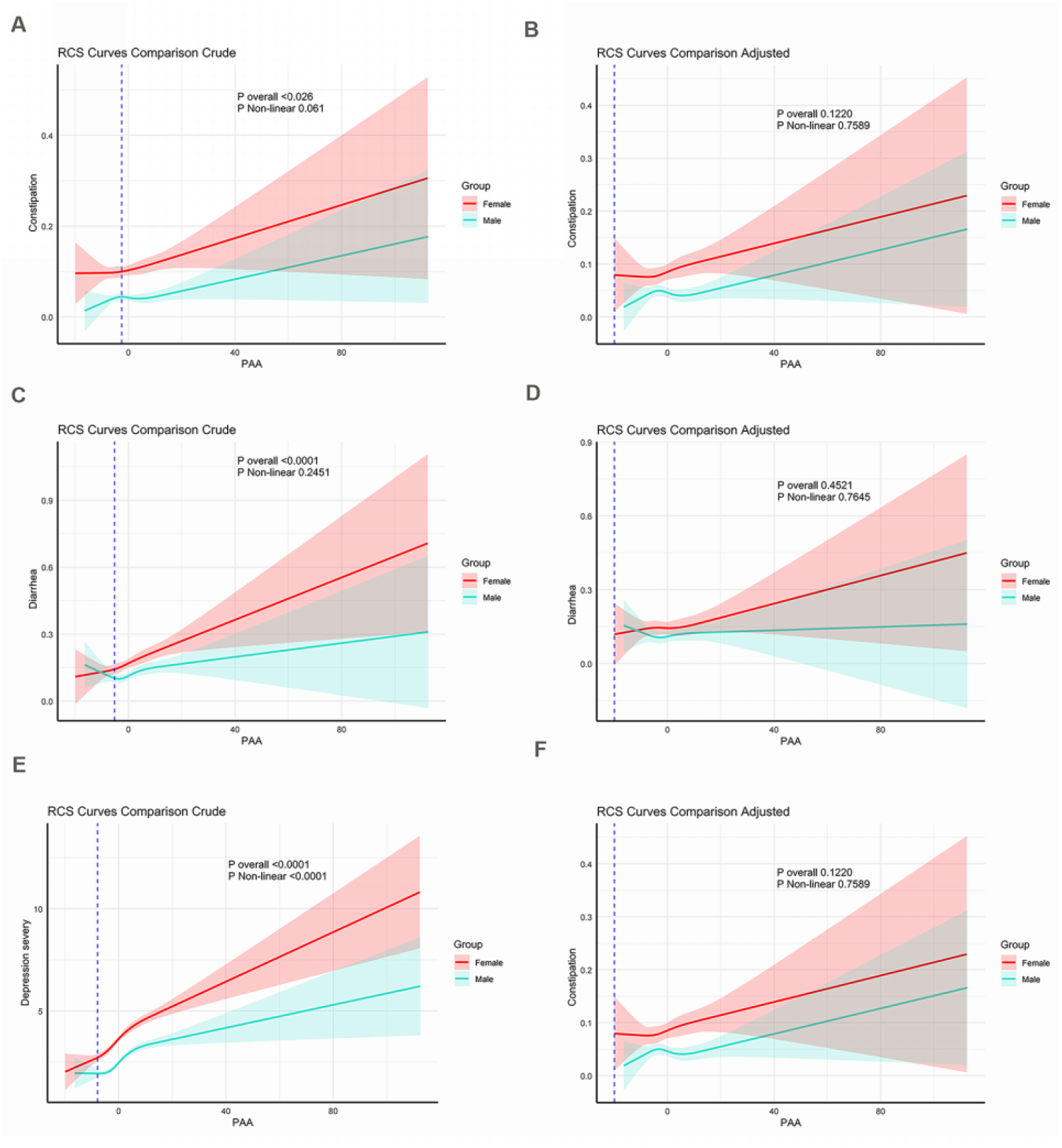
Restricted cubic spline (RCS) analysis of the associations between PAA and the odds of diarrhea, constipation and depression. The figure shows the results of restricted cubic spline analyses with three knots, illustrating the relationship between PAA and the odds of constipation (A, B), diarrhea (C, D) and depression (E, F). The results are presented for both the crude (A, C, E) and fully adjusted models (B, D, F), stratified by sex. The y-axis represents the odds ratio, and the x-axis represents the PAA values. The shaded areas indicate 95 % confidence intervals.

### 3.3. Mediating role of PAA in the relationship between FGIDs and depression severity

To further explore whether PAA exerts a mediating effect between FGIDs and the severity of depression, we first grouped participants according to the diarrhea (Table 3) and constipation (Table 4) subtypes of FGIDs, then constructed mediating effect models for each group respectively, and verified the results through subgroup analysis. The results showed that in the total population of the two subtypes, the mediating effect of PAA was not significant (both P> 0.05); while when subgroup analysis was conducted by age and gender stratification, the two subtypes showed similar trends— the mediating effect of PAA was only significant in participants aged <60 years (both P< 0.05), and the proportion of PAA’s mediating effect between the diarrhea subtype and depression severity was higher (diarrhea group: 4.20924%; constipation group: 7.1004%).

**Table 3.**
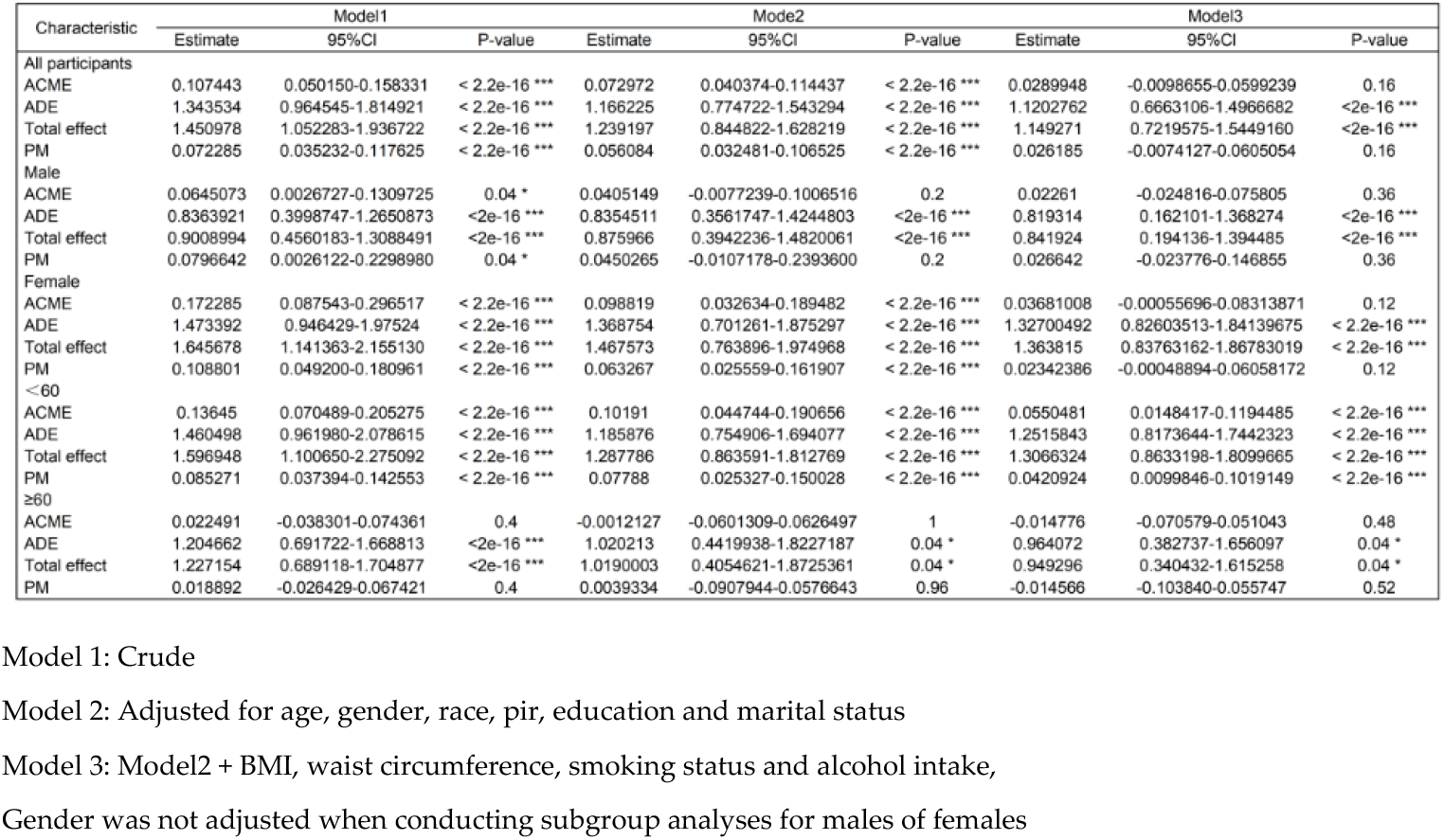
Mediation analyses of the effect of phenotypic age acceleration on the associations between diarrhea and depression severity.

**Table 4.**
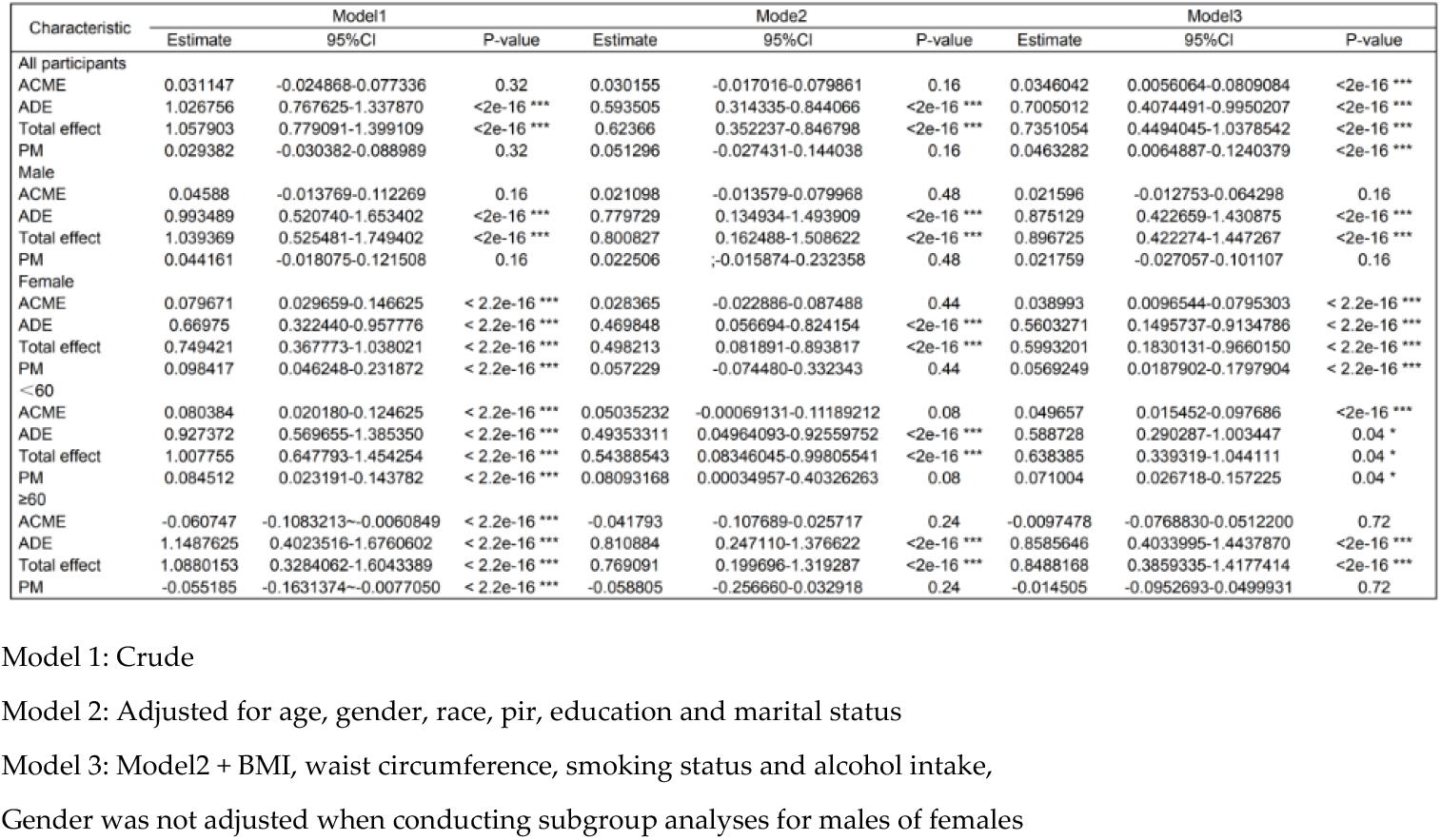
Mediation analyses of the effect of phenotypic age acceleration on the associations between constipation and depression severity.

In gender stratification, among female participants, the mediating effect of PAA was significant when constipation-related variables were not adjusted (diarrhea group: 10.8801%; constipation group: 9.8417%; both P<0.05); after adjusting for variables, only the diarrhea group showed a significant result in the model with adjusted demographic variables (Model 2) (proportion of mediating effect: 6.3267%, P < 0.05), while the constipation group showed a significant result only in the model with all variables adjusted (Model 3) (proportion of mediating effect: 5.69249%, P < 0.05). In addition, among the two subtypes, the mediating effect of PAA was not significant in male participants and participants aged ≥60 years (all P>0.05).

## 4. Discussion

Our study explored whether PAA plays a mediating role in the association between defecation disorders and depression severity. Although previous studies have separately investigated the correlations among these three factors, they were limited to analyzing the associations between only two factors and failed to systematically connect the relationships among the three. Analysis of a nationally representative sample of U.S. adults revealed the following: PAA was positively correlated with both diarrhea and depression severity, while the correlation between PAA and constipation was not statistically significant; results of mediation effect analysis showed that PAA exerted a mediating role in some participants, with the most significant effect observed in participants aged < 60 years, accounting for approximately 4% to 8% of the total effect.

Although the mediating effect was only significant in some populations, it remained reliable after adjusting for relevant variables (P < 0.05). Further subgroup analysis by gender showed that: regardless of whether the independent variable was constipation or diarrhea, the mediating effect of PAA in female participants was statistically significant in the crude model (P < 0.05); when the independent variable was diarrhea, after adjusting for demographic variables, the mediating effect of PAA in the association between diarrhea and depression severity remained statistically significant (accounting for approximately 2% of the total effect, P < 0.05); when the independent variable was constipation, the mediating effect of PAA remained statistically significant in the fully adjusted model (accounting for approximately 5% of the total effect, P < 0.05).

In contrast, among male participants and those aged ≥ 60 years, the mediating effect of PAA was not statistically significant regardless of whether variables were adjusted or not. In conclusion, the results of our study indicate that PAA plays a mediating role in the association between defecation disorders and depression severity, and this mediating role is significantly correlated with age and gender.

Given that constipation and diarrhea are merely a category of clinical symptoms, which can present in diseases such as inflammatory bowel disease (IBD) and irritable bowel syndrome (IBS)[13],[14], we also included such diseases accompanied by constipation or diarrhea in our analysis scope when sorting out previous research findings. The result of “an association between abnormal defecation and depression” observed in this study is highly consistent with the conclusions of prior studies in this field[15],[16]. Existing research has confirmed that IBS and IBD significantly increase the risk of depression, and both affect individuals’ mental health and mental state through three key pathways: chronic inflammatory response, intestinal flora imbalance, and abnormal regulation of the gut-brain axis[17],[18],[19],[20].

Furthermore, this study also found that there are obvious gender differences in the correlation between constipation/diarrhea and the severity of depression: in the linear regression analysis of “constipation and depression severity”, the correlation was higher in male participants (correlation coefficient: 2.33 vs 1.92); while in the correlation analysis of “diarrhea and depression severity”, the correlation was more significant in female participants (correlation coefficient: 3.61 vs 2.39). From the perspective of general population data, the overall correlation between diarrhea and depression was higher than that between constipation and depression (correlation coefficient: 3.09 vs 2.04).

Previous epidemiological studies have shown that there are gender-specific characteristics in the incidence of constipation and diarrhea: in the general population and female groups, the incidence risk of constipation is higher than that of diarrhea; while the incidence characteristics of male groups are the opposite (the incidence risk of diarrhea is higher)[21]. Based on this, we hypothesize that before being included in this survey, females were more likely to have experienced constipation symptoms, and males were more likely to have experienced diarrhea symptoms. With the occurrence of abnormal changes in defecation, participants may have shown an increase in depression scores due to somatic symptom disorder, and the risk of this occurrence was higher in females (correlation coefficient: 3.61 vs 2.33). This result is completely consistent with the conclusions of previous studies that “somatic symptom disorder can predict the risk of depression, and this predictive effect is more significant in females[22]” and “females have a higher risk of depression[23]”. It also further explains why the correlation between diarrhea and depression is higher than that between constipation and depression in the general population.

Furthermore, the link between depression and accelerated aging has been gradually confirmed. Prospective studies have shown that accelerated biological aging, particularly accelerated phenotypic age, is significantly associated not only with the presence of depression at baseline but also with the risk of developing new-onset depression in the future[24]. This finding is consistent with our observation in populations with abnormal bowel movements, where we also found a correlation between accelerated phenotypic age and the severity of depression.

Another exploration focus of this study was to investigate whether PAA plays a mediating role in the correlation between constipation (or diarrhea) and depression severity. The results revealed that in individuals aged < 60 years, PAA significantly mediated the association between constipation (or diarrhea) and depression severity. This mediating effect remained statistically significant even after adjusting for relevant variables (in the constipation group, the mediating effect accounted for approximately 7%; in the diarrhea group, it accounted for approximately 4%, with all P-values < 0.05). However, no reliable evidence of PAA’s mediating role was observed in the subgroups of participants aged ≥ 60 years or male participants in either group. In the female subgroups of both groups, the mediating effect of PAA was only statistically significant in the unadjusted crude model and some partially adjusted models. From an epidemiological perspective, this result may be related to the incidence characteristics of IBS, IBD, and depression—all three diseases show higher incidence rates in female populations and individuals aged < 60 years[25],[26],[27],[28].

Previous studies have confirmed that the persistence of constipation or diarrhea affects overall health through mechanisms such as intestinal flora imbalance and chronic inflammation[29],[30],[31], and these mechanisms overlap with the changes that occur in the body during the aging process. For instance, aging is accompanied by alterations in the composition of intestinal flora: in the elderly population, the number of beneficial bacteria (e.g., *Coprococcus, Faecalibacterium, Lactobacillus*) in the intestines decreases, while the number of pro-inflammatory “opportunistic pathogens” (e.g., *Fusobacterium, Staphylococcus*) increases. This flora imbalance impairs the integrity of the intestinal barrier and reduces the production of short-chain fatty acids (SCFAs)—substances that possess anti-inflammatory properties and are crucial for maintaining intestinal barrier function[32],[33]. Additionally, a growing body of evidence indicates an association between mental health and aging; depression can accelerate the aging process by inducing chronic inflammation, oxidative stress, hormonal imbalances, and other pathways[34]. The aforementioned research findings provide a theoretical basis for explaining why PAA can act as a mediating variable in the correlation between abnormal bowel movements and depression.

This strengths of our study are as follow. First, the NHANES database used in the study not only has a large sample size and a long survey period but also is well-representative of the U.S. population, laying a solid foundation for the reliability of the research results. Second, this study adopted Phenotypic Age Acceleration (PAA) to assess aging status, which covers the functions of multiple systems throughout the body and is a well-recognized and reliable indicator in aging-related research. Third, through mediation analysis, this study provides new research perspectives and insights into the association mechanism among abnormal bowel movements, aging, and depression.

However, our study also has several limitations. Firstly, the cross-sectional design of the study only allows the analysis of correlations between variables, but cannot determine the causal relationships among the three. Secondly, due to the lack of longitudinal follow-up data, it is impossible to further evaluate the long-term dynamic changes and effects of the above-mentioned correlations. Thirdly, the diagnosis of constipation and diarrhea in participants relies on a questionnaire designed based on the Bristol Stool Scale. This method may lead to recall bias among participants, which affects the accuracy of the research results. Finally, the study participants are mainly from the general population of the United States, and this population characteristic may, to some extent, limit the generalization of the research results to populations in other countries or regions, reducing the universality of the results.

## 5. Conclusion

In conclusion, this study demonstrates that individuals with abnormal bowel movements exhibit accelerated aging, specifically manifested by an increase in PAA. Moreover, in this population, PAA may play a mediating role in the association between abnormal bowel movements and the severity of depression. These findings emphasize that interventions for individuals with abnormal bowel movements should simultaneously focus on their mental health status, while attaching importance to the impact of biological aging processes in this context. Future research should further explore whether interventions targeting aging-related processes can provide new directions for the treatment of abnormal bowel movements and depression. Additionally, in-depth investigation into the molecular mechanisms underlying the aforementioned associations may not only offer deeper insights into the pathogenesis of both abnormal bowel movements and depression but also is expected to promote the development of novel therapeutic strategies.

## Abbreviations

PAA: phenotypic age acceleration
PA: phenotypic age
PHQ-9: Patient Health Questionnaire-9
RCS: Restricted Cubic Spline
CI: Confidence Interval
NHANES: National Health and Nutrition Examination Survey
FGIDs: functional gastrointestinal disorders
PIR: poverty income ratio

## Declarations

None

## Acknowledgments

The authors appreciate the technical help from staff in the Department of General Surgery at the General Hospital of Northern Theater Command.

## Funding

This study was supported by Liaoning province livelihood science and technology joint plan (No. 2024JH2/102600293).

## Author contributions

GHZ, XNW, XYW and CZ contributed to the conception and design of this study. GHZ and XNW performed the statistical analysis. This first draft of the manuscript was written by GHZ, and all authors commented on the previous versions of the manuscript. All authors have read and approved the final manuscript.

## Conflict of Interest

The authors declare no competing interests.

## Data Availability

The data supporting the findings of this study are publicly available from the National Health and Nutrition Examination Survey (NHANES) website, managed by the National Center for Health Statistics (NCHS). Data can be accessed at: https://www.cdc.gov/nchs/nhanes/?CDC_AAref_Val=https://www.cdc.gov/nchs/nhanes/index.htm

## Consent for publication

not applicable.

## Clinical Trial Number in the manuscript

not applicable.

## Ethics approval and consent to participate

not applicable.

## Notes

### Competing Interest Statement

The authors have declared no competing interest.

